# Mobilizing faith-based COVID-19 health ambassadors to address COVID-19 health disparities among African American older adults in under-resourced communities: A hybrid, community-based participatory intervention

**DOI:** 10.1101/2023.05.07.23289636

**Authors:** Edward K. Adinkrah, Shahrzad Bazargan, Sharon Cobb, Lucy W. Kibe, Roberto Vargas, Joe Waller, Humberto Sanchez, Mohsen Bazargan

**Affiliations:** Department of Family Medicine, Charles R. Drew University of Medicine and Science, Los Angeles, CA, USA; Departments of Psychiatry, Charles R. Drew University of Medicine and Science, Los Angeles, CA, USA; Department of Psychiatry and Biobehavioral Sciences, University of California Los Angeles, CA, USA; Mervyn M. Dymally College of Nursing, Charles R. Drew University of Medicine and Science, Los Angeles, CA, USA; Physician Associate Program, Charles R. Drew University of Medicine and Science, Los Angeles, CA, USA; Department of Internal Medicine, Charles R. Drew University of Medicine and Science, Los Angeles, CA, USA; Office of Research, Charles R. Drew University of Medicine and Science, Los Angeles, CA, USA; Department of Family Medicine, University of California Los Angeles, Los Angeles, CA, USA

**Author notes:** Corresponding author (E.K.A).

## Abstract

The COVID-19 pandemic disproportionately affected older adults, particularly those with pre-existing chronic health conditions. To address the health disparity gap and challenges faced by under-resourced African American older adults in South Los Angeles during this period, we implemented a hybrid (virtual/in-person), pre-post, community-based participatory intervention research project utilizing a faith-based lay health advisor model (COVID-19 Health Ambassador Program (CHAP)). We recruited COVID-19 Health Ambassadors (CHAs) and African American older adults (participants) from faith-based organizations who partook in CHA-led meetings and follow-ups that educated and supported the participants. This paper seeks to evaluate this intervention’s implementation using the Consolidated Framework for Implementation Research (CFIR) as a reporting tool with an emphasis on fidelity, challenges, and adaptations based on data collected via stakeholder interviews and surveys. Results: CHAP was delivered to 152 participants by 19 CHAs from 17 faith-based organizations. CHAs assisted with chronic disease management, resolved medication-related challenges, encouraged COVID-19 vaccination, reduced psychological stress and addressed healthcare avoidance behaviors such as COVID-19 vaccine hesitancy among the participants. Challenges encountered include ensuring participant engagement and retention in the virtual format and addressing technological barriers for CHAs and participants. Adaptations made to better suit the needs of participants included providing communication tools and additional training to CHAs to improve their proficiency in using virtual platforms in addition to adapting scientific/educational materials to suit our participants’ diverse cultural and linguistic needs. Conclusion: The community-centered hybrid approach in addition to our partnership with faith-based organizations and their respective COVID-19 health ambassadors proved to be essential in assisting underserved African American older adults manage chronic health conditions and address community-wide health disparities during the COVID-19 pandemic. Adaptability, cultural sensitivity, and teamwork are key to implementing health interventions especially in underserved populations.

## Introduction

The COVID-19 pandemic posed significant challenges in managing chronic health conditions, particularly among underserved African American (AA) older adults with underlying health issues (1–3). This under-resourced population, who have historically relied on county-based safety-net facilities for their healthcare needs(4), were forced to adjust their health-seeking behaviors and patterns of necessary medical care to manage their chronic health conditions. Consequently, this may have resulted in delayed, reduced, or halted visits to primary and specialty healthcare providers or pharmacies for medication (3). Moreover, pre-existing health disparities and conditions may have been exacerbated due to limited access with healthcare providers/resources and medication availability and adoption of risky health behaviors, including non-adherence to chronic health condition management guidelines, unhealthy lifestyles, and dietary practices (5). In response to these challenges and the urgent need for timely and health information during the pandemic(6), innovative approaches were necessitated to address health disparities and promote effective self-management of chronic health conditions.

This particular approach capitalizes on the external influence of trusted community organizations and leaders, such as faith-based organizations (FBOs) who have been historically recognized as vital sources of social support and resources for AA communities (7). More importantly, the engagement of Lay Health Advisors (LHAs) (8) from these FBOs are recognized as community liaisons trained to provide health-related support, guidance, and education to their peers (9). By sharing cultural, linguistic, and socioeconomic backgrounds with the populations they serve, LHAs foster trust, rapport, and understanding (10). The integration of LHAs from community churches into health interventions has demonstrated effectiveness in improving health outcomes and addressing the unique needs of AA older adults with chronic health conditions (11–13).

To adapt to the unique challenges of the COVID-19 pandemic, including the necessity for social distancing and reduced face-to-face interactions, a community-based participatory intervention utilizing a hybrid LHA model, the COVID-19 Health Ambassador Program (CHAP), was implemented. This model combined the benefits of in-person and virtual/telephone interactions, leveraging internet-enabled devices/phones and the growing digital literacy among older adults to facilitate remote support and engagement while maintaining the essential personal connection (14, 15). This paper aims to evaluate the successful implementation of CHAP in addressing health disparities among underserved AA older adults during the COVID-19 pandemic using the Consolidated Framework for Implementation Research (CFIR) as a reporting tool.

## Methods and materials

This study utilized a mixed-methods, community-based participatory research (CBPR) approach to implement a “one group, pretest–posttest” intervention over a two-year period. All participants provided written, informed consent before participating, and the study protocol was approved by the ethical committee of the Charles R. Drew University of Medicine and Science Institutional Review Board (IRB). Authors had no access to information that could identify individual participants during or after data collection.

### Setting

This study took place in 17 predominantly African American FBOs primarily located in urban regions of Los Angeles County Service Planning Area (SPA) 6, which experienced a significant impact from the COVID-19 pandemic. Compared to the rest of Los Angeles County, SPA 6 individuals face increased health challenges at a disproportionate rate.(16) The FBOs, situated within a 10-mile radius of the primary study site, had a well-established membership of older adults who frequently participated in religious services and maintained ongoing relationships with their respective leadership (e.g. pastors, ministers, deacons, etc.).

### Recruitment

The study’s lead community faculty (CF), who is also serves as the head pastor of a participating FBOs, initiated contact with other head pastors (HP) and prominent faith-based leaders (FL) to encourage their participation in the research project. Interested FBOs with their respective leaders were conveniently sampled and recruited into the study between 2020 and 2022. No FBO was excluded based on denomination or size of the congregation.

Potential CHAs were conveniently sampled from our 17 partner FBOs. Based on recommendations from individual faith-based leaders, trusted parishioners who voluntarily signed up to become CHAs were chosen based on the following criteria: 1) 18 years and older, 2) AA parishioner from a registered partner FBO, 3) attend a 3-day workshop training, 4) provide at least 3 hours per week for study activities, 5) strongly committed to assisting older adults with chronic illness management and COVID-19 risk reduction, 6) familiar with AA community, and 7) able to communicate effectively in both English and the preferred language of the older adults.

Also, the FBOs provided access to potential participants of the study. These would be members who were 65 years and older or 55 years and older with at least one chronic health condition. Residents in care facilities and those with cognitive deficits (identified by the short version of the mini-mental state examination instrument) were excluded from the intervention.

### Data collection

Data on participant and CHA sociodemographic characteristics, chronic health conditions, and health status were gathered through surveys. Participants completed the study surveys using different methods, including Uniform Resource Locator (URL), telephone interview, or CHA-administered interview. Additionally, participants’ acceptance and completion rates throughout the intervention were assessed. Complementing the survey-based approach, in-depth interviews were employed to gain a deeper understanding of the FBO leaders’ and CHAs’ viewpoints regarding the project and its implementation process. As part of the data-gathering process, CHAs and older adult participants identified and prioritized significant barriers and facilitators to implementing the intervention within their church or community setting.

### Analysis plan

Data analysis was conducted using descriptive statistics for the quantitative data and thematic analysis for the qualitative data. CFIR, a useful tool for assessing potential barriers and facilitators in implementing healthcare interventions, was utilized to guide the analysis of the implementation process and identify areas for improvement. This tool provides a practical, theory-based guide to tailor implementation strategies and adaptations based on these factors and explain the outcomes of the implementation process(17). The project team also documented any adaptations made to the intervention based on feedback from the CHAs and older adults. The participants’ identified facilitators and barriers were grouped and reviewed before being compiled. The quantitative data were analyzed using SPSS (version 25). To produce a narrative report based on the CFIR domains, qualitative data were organized according to the CFIR constructs and sub-constructs.

### Implementation strategies

Our project employed innovative implementation strategies by leveraging the trust and expertise of all stakeholders. These formed the basis of the project’s conceptual model and broadly described below.

### Community strategizing

A six-step strategy was developed to forge strong relationships with the FBO leadership and adapt COVID-19 public health guidelines for their respective denominations. This approach involved recruiting leaders, creating a specialized curriculum, and conducting a 3-day virtual training workshop to empower them in addressing COVID-19-related concerns within their communities. FBO leaders were responsible for implementing infection prevention measures, carrying out routine environmental assessments, and ensuring protocol compliance. Additionally, they participated in community outreach through the university’s weekly public radio show that promoted the CHAP’s goals and discussed culturally sensitive topics during the pandemic. These included, 1) ‘Managing COVID-19 Grief in Our Community’, 2) ‘Mental Health and Social Isolation among African Americans’, 3) ‘How to Mitigate Vaccine Barriers in our Communities’, 4) ‘How ‘Your Health Is Connected to Your Faith’, and 5) ‘Addressing Vaccine Hesitancy among the Youth; The Role of the Church’. FBOs supported the project by recruiting lay health advisors, coordinating communication between advisors and participants, and facilitating participant involvement.

### Community building and sustainability

The individual-level collaborations established with FBO leadership enabled the development of a tailored CHA curriculum and the dissemination of the CHA workshop/training program. The courses within the workshop covered CHA roles, COVID-19 knowledge, testing and vaccination concerns, risk prevention strategies, social determinants, chronic disease management, available community resources, and specialized care delivery (Table 1). The 3-day virtual workshop was facilitated by FBOs, research/academic staff, healthcare providers, and community faculty, with recorded sessions available to those who missed the live event and for CHAs who needed refresher training. CHAs received iPads and were trained in confidentiality and data security.

**Table 1.** List of Topics and Presenters as Used in Training Guideline for CHAs

| Topics Covered | Presenter |
| --- | --- |
| Functions and expectations of lay health advisors | Research Staff/Church Leader |
| Medical & Research Mistrust | Public Health Specialist |
| COVID-19 testing and vaccination in community settings | Physician 1 |
| COVID-19 vaccine equity | Physician 2 |
| Mental health and COVID-19 related trauma | Mental Health Nurse |
| Chronic disease management (with and without COVID-19 diagnosis) | Physician |
| Telehealth and online services | Physician 3 |
| How to be a church-based health advisor | Public Health Specialist<br>/Church Leader 1 |
| Motivational interviewing in the church | Church Leader 2 |
| How to communicate with parishioners on their medications | Pharmacist 1 |
| Alternative treatments to COVID-19 | Pharmacist 1 |
| Linking community resources to community needs | Community Faculty |
| Health tracking and data collection | Research Staff |
| Immunization | Public Health Nurse |

**Table 2.** Recruitment and Completion Rates of CHAs and Participants

|  | Cohort One | Cohort Two | Total |
| --- | --- | --- | --- |
| **Faith-Based Organizations** |  |  |  |
| Participating Churches | 8 | 9 | 17 |
| **Faith-Based COVID-19 Ambassadors** |  |  |  |
| Prospective Number of COVID-19 Ambassadors<br>(three per church) | 24 | 27 | 51 |
| Trained COVID-19 Ambassadors | 24 | 23 | 47 |
| Active COVID-19 Ambassadors during Intervention | 14 | 5 | 19 |
| Participation rate of Trained COVID-19 Ambassadors<br>(trained/expected number) | 100% | 85% | 92% |
| Participation rate of Active COVID-19 Ambassadors<br>(active/trained) | 58% | 22% | 40% |
| **African American Older Adult Parishioners**<br>**(Participants targeted for CHAP)** |  |  |  |
| Number of prospective participants (10 participants<br>from each active ambassador) | 140 | 50 | 190 |
| Number of recruited participants pre-CHAP<br>(completed baseline survey) | 118 | 34 | 152 |
| Number of participants who completed CHAP | 90 | 20 | 110 |
| Percentage of participant completed CHAP<br>(participants recruited pre-CHAP/participants who<br>completed CHAP) | 76% | 67% | 72% |

**Table 3.** Sociodemographic characteristics of Participants and CHAs

| Sociodemographic Characteristics | Participants Targeted for CHAP (Older African American Parishioners), N=152, N (%) | CHAs, N=19, N (%) |
| --- | --- | --- |
| **Gender** |  |  |
| Male | 45 (30) | 3 (16) |
| Female | 105 (70) | 16 (84) |
| **Age** |  |  |
| 40-54 | 0 (0) | 1(5) |
| 55-64 | 48 (32) | 7 (37) |
| 65-74 | 68 (45) | 10 (53) |
| 75 and older | 34 (23) | 1 (5) |
| **Education** |  |  |
| No High School Diploma | 19 (13) | 0 (0) |
| High School Diploma | 40 (27) | 2 (27) |
| Some college/graduate | 91 (60) | 17 (73) |

### Individualized management of care

The project team collaborated to create culturally appropriate intervention materials and trained CHAs to support participants via phone calls and videoconferencing remotely. The three-month intervention included five broad activities: 1) completion of pre-and post-intervention surveys, 2) attendance at six bi-weekly online/hybrid meetings, 3) participation in individualized checkups/visits, 4) project support and feedback, and 5) participation in a post-study conference/gala. The team coordinated with FBOs to align schedules and ensure CHA availability. CHAs helped older adults develop personalized action plans for managing chronic health conditions, addressing medication management, healthy eating, and physical activities.

## Results

The intervention engaged 17 AA FBOs in South Los Angeles, with additional churches in Southern California’s Inland Empire, Antelope Valley, and Victorville (n=3). On a daily average, 47 CHAs from these FBOs attended the 3-day workshop, and 19 CHAs actively participated in the recruitment and support of participants. Each CHA aimed to recruit 10 participants, resulting in a 22% rejection rate, as 2 out of 10 parishioners averagely declined the 19 CHAs’ invitations to participate in the study’s baseline data collection. Consequently, out of the 152 participants who enrolled in the study, 110 participants completed the intervention, yielding a 72% completion rate.

The majority of participants (70%) were female, with a mean age of 69 (SD: 9), and 25% were aged ≥75 years. Approximately 14% did not complete high school, 33% lived alone, and 98% had health insurance. Over 35% reported poor or fair physical health, and participants had an average of two chronic health conditions. The most prevalent comorbidities were hypertension (59%), COPD or asthma (24%), diabetes mellitus (22%), and heart disease (11%).

The implementation process was evaluated through a combination of informal interviews with FBO leadership, CHAs and participant survey items. Based on CFIR reporting guidelines, the findings highlighted the intervention characteristics (five constructs), inner setting (three constructs, three subconstructs), and the process (three constructs, two sub-constructs) as the most frequently addressed components of the CFIR, while individual characteristics (one construct) received comparatively less attention in the data. This is likely due to the intervention’s focus on the FBOs and the CHAs collectively rather than individually. The influential components of the CFIR that emerged from the data are illustrated in Fig 2.

**Fig 1.**
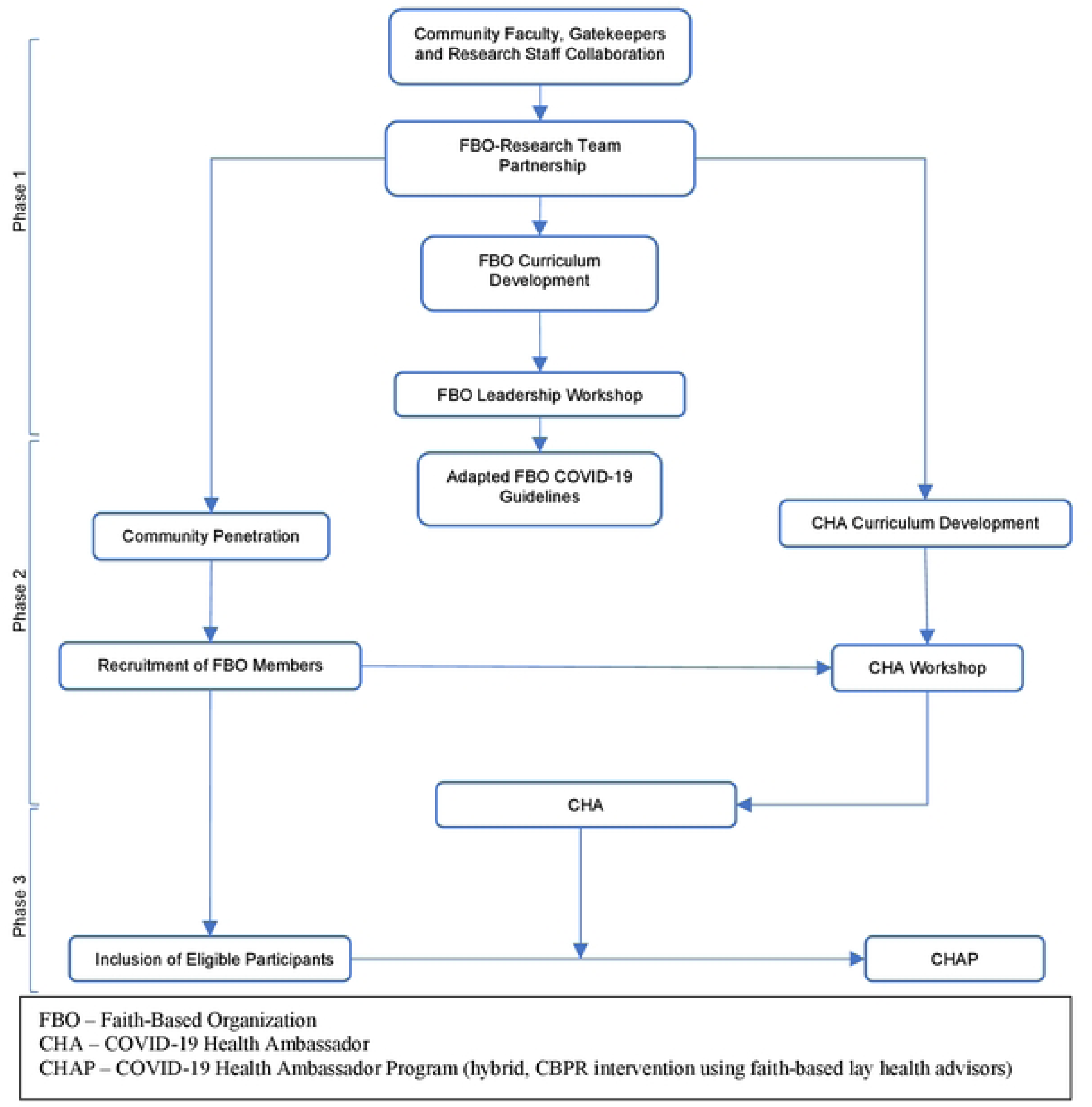
Conceptual model of a hybrid community-partnered intervention project using faith based lay health advisors.

**Fig 2.**
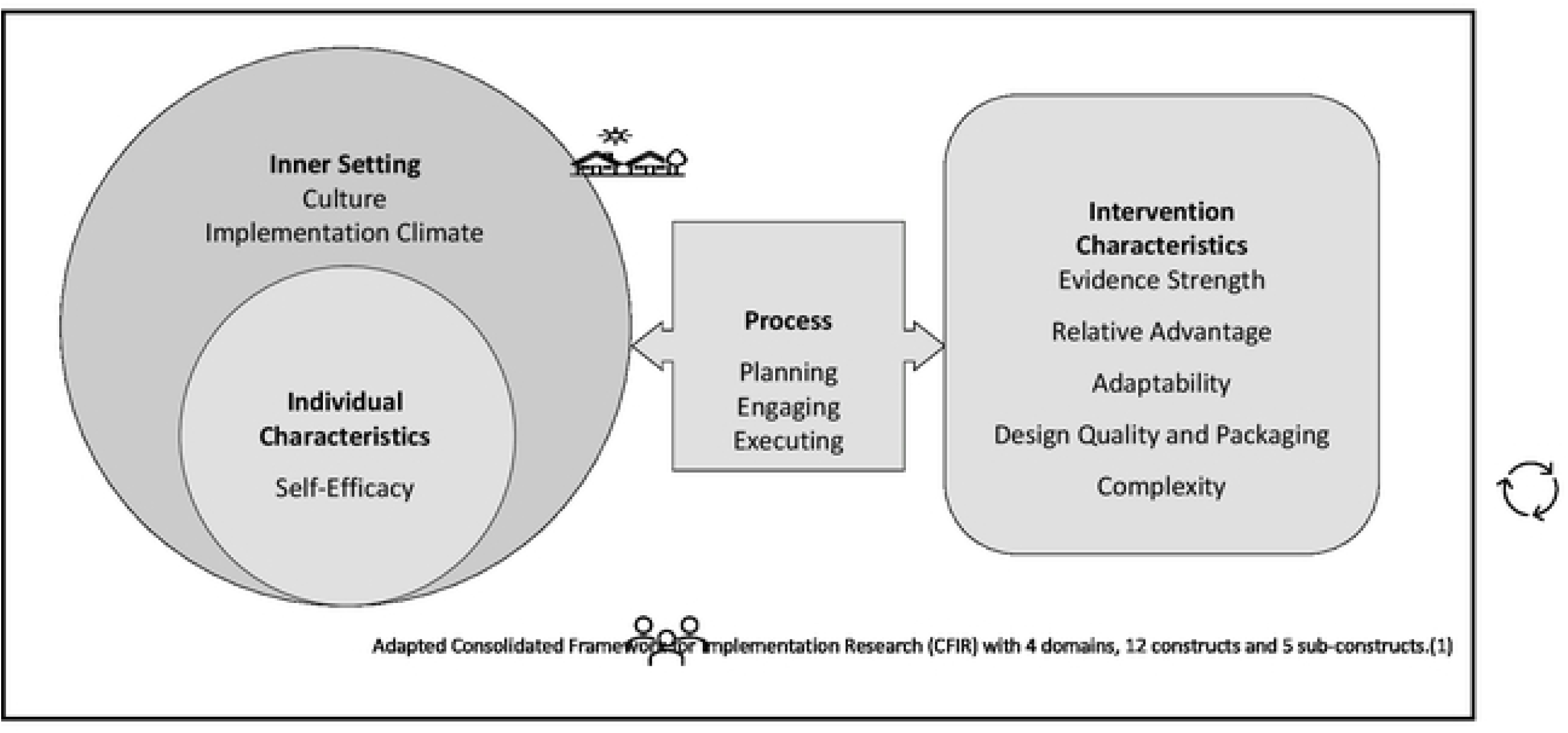
Adapted Consolidated Framework

### CFIR-related intervention characteristics

#### a) Evidence strength

The project utilized a CBPR approach, considered a high-quality study design in the field of public health research (18). The project’s use of faith-based health advisors aligns with evidence supporting this approach’s effectiveness in closing unmet needs gaps and decreasing health disparities for minority communities, including AAs (11, 12, 19–23). The CDC “scaling up operation” recommends that faith-based organizations (FBOs) establish and maintain communication with local authorities and work closely with health organizations to implement COVID-19 guidelines Field [22] effectively. Telehealth methods had been suggested as an innovative approach to delivering healthcare services to vulnerable populations, such as AAs, who experience disparities in accessing and maintaining healthcare services (24–26). AA churches have historically supported their families economically, spiritually, socially, and culturally through programs that are unmatched by other social institutions (11).

#### b) Relative advantage

CHAs reported various advantages of telehealth technology, including improved access to primary care provider, reduced barriers to support, and more frequent communication with participants. This allowed for personalized and timely assistance, reduced in-person visits during the pandemic, and provided greater flexibility in scheduling provider appointments.

#### c) Adaptability

The COVID-19 pandemic greatly impacted chronic disease management for underserved older adults, with social distancing limiting in-person visits. Telehealth was initially used but faced technological barriers and rapport challenges. The project team adapted with a hybrid model, combining in-person and telehealth check-ins, allowing for personalized care and improved communication.

#### d) Design quality and packaging

The research project ensured design quality by providing comprehensive training to FBO leaders and CHAs through a 3-day virtual workshop. FBO leaders received computers, while CHAs were given iPads, resource manuals, and biweekly discussion topics with detailed links to data and resource materials. Continuous feedback and response mechanisms were implemented through weekly meetings with research staff.

#### e) Complexity

The project involved bi-weekly meetings for nine months, with maintaining regular communication between CHAs and older adults. Pre-and post-intervention surveys focused on nutrition, physical activity, food insecurity and a health tracking assessment was utilized to measure progress. A hybrid model of videoconferencing, phone calls, and in-person sessions provided flexibility, but also required reliable internet and access to technology. The team addressed these challenges by offering technical support and troubleshooting.

### CFIR-related inner settings

The research team implemented a comprehensive communication plan promoting regular stakeholder engagement, including virtual weekly check-ins between research staff and CHAs. The approach enabled collaboration and innovation by fostering strong relationships between CHAs and the older adults they served and facilitating peer-support groups and team-building activities among CHAs, which proved crucial during the COVID-19 pandemic.

#### a) Culture

The use of CHAs from the churches also ensured that the project was culturally appropriate and responsive to the community’s needs. Several specific examples of the culture construct can be highlighted to demonstrate how it was addressed and integrated throughout this project’s implementation process. Some included:

> <u>Cultural beliefs about health and illness</u> – The project addressed community preferences for traditional or alternative medicine by training CHAs to integrate these beliefs into evidence-based self-management strategies.

> *“As a CHA, I encountered an older adult who relied on herbal remedies to manage their diabetes. I worked with them to ensure these remedies were used alongside conventional treatments, resulting in better blood sugar control.”* (FBO#1,CHA#2)

> <u>Dietary practices and preferences</u> – The research team ensured that culturally relevant nutritional advice was provided, considering specific dietary practices and preferences within the AA community.

> *“One of my older adults loved soul food, so I helped them find ways to prepare their favorite dishes using healthier ingredients and cooking methods, making their meals both culturally satisfying and beneficial to their health.”* (FBO#2,CHA#1)

> <u>Language and communication</u> – CHAs were recruited for their familiarity with the cultural context and communication styles of AA older adults, and educational materials were adapted to be culturally appropriate.

> *“We made sure our educational materials spoke to our community, using analogies and references that our older adults could easily relate to. This approach made complex health concepts more accessible and understandable.”* (FBO#4,CHA#2)

> <u>Family and community dynamics</u> – CHAs were encouraged to engage with the older adults, their families, and community networks, fostering a supportive environment for self-management strategies.

> *‘Most of the people I recruited were my family – my husband and sisters. Going through this process, I was able to find out about everyone’s health issues that I never knew they dealt with in the past.’* (FBO#1,CHA#1)

> <u>Traditional community gatherings</u> – The research team used traditional AA community gatherings as opportunities to raise awareness about the health ambassador program and engage with the community, demonstrating respect for the community’s culture and building trust with older adults and their families.

> *“I used our weekly dinner and bingo sessions to talk with community members about the health ambassador program. We shared stories, addressed concerns, and offered support in a relaxed, familiar environment. It helped us build trust and connect with the community on a deeper level.”* (FBO#5,CHA#3)

#### b) Implementation climate

The implementation climate for the CHA intervention required a sense of shared commitment, necessary resources, training, and clear goals. Issues related to clarity posed challenges, including confusion about objectives, roles, and expected outcomes. To address these issues, the research team organized additional meetings and workshops, developed detailed guidelines, established more explicit performance metrics, and implemented regular check-ins with CHAs and community partners. These efforts helped maintain clarity, resolve emerging issues, and reinforce the shared commitment to the program’s goals.

> <u>Tension for change</u>: This awareness of existing health disparities within our target communities created a sense of urgency and motivation for change among the research team, CHAs, and community partners, fostering a conducive environment for implementing the CHA intervention. The lack of support felt by AA older adults, heightened concern, misinformation, disinformation, confusion messaging and awareness of the risks associated with the pandemic increased this tension for change as stakeholders recognized the need for additional support and resources.

> As the CHA intervention was implemented, the research team gathered success stories from older adults who had benefitted from the support provided by the CHAs. Sharing these success stories with the broader community, including those who were initially resistant to the program, helped demonstrate the intervention’s positive impact and increased the tension for change by showcasing the program’s benefits.

> <u>Compatibility</u>: To address the specific needs of each participant, the project permitted CHAs to customize the delivery of interventions based on the choices and circumstances of each older adult (*Kangovi et al., 2014*). Depending on the participant’s comfort level and accessibility, CHAs selected in-person visits, phone conversations, or virtual meetings. Its adaptability boosted the program’s compatibility with the various demands of the older adults and aided in sustaining participation throughout the intervention.

> <u>Relative Priority:</u> Non-participating local church leaders and FBOs collaborated with the project team, referring older adults needing CHA support and actively participating in evaluating and improving the intervention.

### CFIR-related individual characteristics

#### Self-efficacy

The project equipped CHAs with the knowledge, skills, and resources to provide tailored support, broadly classifying the approach under training, social support, and peer modeling.

> Training: CHAs were trained to employ goal-setting, problem-solving, and self-monitoring approaches to assist older persons in gaining confidence in their abilities to manage chronic diseases and stick to treatment programs.

> Peer Modeling: The CHAs, who shared comparable cultural and socioeconomic backgrounds with the older persons they supported, functioned as role models by displaying good self-management skills and highlighting success stories from their communities. Peer modeling played a crucial impact in boosting older individuals’ self-efficacy.

> CHAs offered personal experiences or stories of community members who effectively treated their chronic health conditions, demonstrating the viability of adopting healthy practices and highlighting the advantages of active self-management.

> Social Support: The CHAs also gave older persons with social support, boosting their self-efficacy by establishing a sense of belonging and connection. This assistance comprised emotional encouragement, educational support, and practical solutions in managing their chronic diseases. CHAs provided reassurance, answered concerns, and assisted with chores such as organizing medical visits and navigating the healthcare system.

### CFIR-related process

#### a) Planning

The project team began by clearly outlining the objectives and goals of the intervention, such as improving chronic disease management among participants and increasing their engagement in health-promoting behaviors. Next, the team conducted a thorough assessment of the available resources, including human and technological resources, to support the implementation of the intervention. For instance, we identified local health professionals within the churches who could serve as CHAs and ensured access to necessary technological tools. After assessing the resources, the project team developed a detailed timeline for the implementation of the intervention, which included milestones and deadlines for each phase of the project (Table 4).

**Table 4.** Project Timeline

| Activity | Months |  |  |  |  |  |
| --- | --- | --- | --- | --- | --- | --- |
|  | 1-3 | 4-6 | 7-9 | 10- 14 | 15-19 | 20-24 |
| Recruitment of FBOs | x |  |  |  |  |  |
| FBO curriculum development and workshops | x |  |  |  |  |  |
| Recruitment and training of FBO lay health advisors | x | x |  |  |  |  |
| Recruitment of participants |  | x | x |  |  |  |
| Collection of baseline data from participants |  |  | x |  |  |  |
| Implementation of CHAP |  |  |  | x | x |  |
| Collection of post-CHAP data from participants and CHAs |  |  |  |  |  | x |

Lastly, recognizing that unforeseen challenges might arise during the implementation of the intervention, particularly given the uncertainty of the COVID-19 pandemic, the project team developed contingency plans to address potential barriers. For instance, we prepared alternative training methods and modes of communication to accommodate potential lockdowns or changes in public health guidelines.

#### b) Engaging

Ten out of the seventeen FBOs representing the study’s first cohort with their respective leaders actively partook in the study. Each participating FBO leader was sent an invitation notice via email, which contained information regarding the study. Once an expression of interest to participate was received, the research team met with these leaders to deliberate the expectations and research plan. FBO leaders were crucial in recruiting potential volunteers as CHAs based on their experience, interest, and ability to work with older adults in their congregation. FBOs held virtual meetings to discuss the project’s goals and objectives with interested volunteers and provided them with details of the eligibility criteria. They also used various communication channels such as bulletins, newsletters, and announcements during the few in-person services to spread the word about volunteer opportunities.

> <u>Opinion leaders (Formally appointed implementation leaders)</u>: As internal implementation leaders, FBO leaders faced competing priorities and interests, making it challenging to consistently support the project. The research team implemented strategies such as flexible scheduling, frequent communication, and progress updates to address this. The research team maintained their support and involvement by accommodating the FBO leaders’ busy schedules.

> <u>Champions:</u> CHAs faced challenges balancing responsibilities and staying up-to-date with relevant information. The research team supported CHAs through refresher training, providing iPads, offering recognition, weekly check-ins, and maintaining constant communication. By addressing these challenges and offering flexible scheduling, the team created an environment for CHAs to thrive.

#### c) Executing

To enhance CHA recruitment efforts by leaders of various FBOs, the research team employed a combination of strategies and procedures to ensure we attracted and retained dedicated, competent and diverse group of individuals. These strategies and procedures included:

> External Collaborations: The research team partnered with local community organizations, such as African American Community Empowerment Council (AACEC), to identify potential CHAs within their target population. This approach facilitated the recruitment of individuals who were already active in their communities and had established connections with the older adults they would support.

> Word-of-mouth referrals: The research team encouraged existing CHAs and other community members involved in the project to refer potential candidates from their personal and professional networks. This strategy helped identify individuals who were passionate about helping others and had the interpersonal skills necessary to succeed as a CHA.

> Social Media: The research team utilized various advertising channels, such as our FBOs’ social media platforms and radio announcements, to reach a wider audience and inform them about the opportunity to become a CHA. This strategy increased the visibility of the project and attracted a diverse pool of candidates.

## Discussion

Our findings from 17 AA churches in South Los Angeles revealed a high project completion rate (72%) among participants. This success can be attributed to the implementation of a culturally sensitive, contextually relevant, and responsive approach achieved through CBPR. This approach, which involved partnering with community churches, likely increased participants’ trust and engagement with the intervention. (18, 27, 28). These findings align with existing literature on community-based health interventions targeting underserved populations. A study by Kangovi and colleagues (2014) implemented a community health worker intervention in a low-income urban population and observed significant improvements in chronic disease control and mental health outcomes (29). Similarly, Resnicow and colleagues (2006) found that a culturally tailored intervention was more effective in achieving behavior change among AA adults attempting to increase fruit and vegetable consumption (30).

We also observed a considerably higher (70%) number of female participants in this study. In comparison, Kangovi and colleagues (2014) also observed a larger number of female participants (69%) (29). Women, particularly in AA communities, often play a vital role in the social and spiritual life of their communities, with churches traditionally serving as the focal point (7). Additionally, women tend to be more health-conscious and are more likely than men to participate in preventive health initiatives(31). They are also more inclined to seek medical care, adhere to medical advice, and engage in health-promoting behaviors (32). CHAP’s focus on underserved AA older adults with chronic health conditions is consistent with other research highlighting the need for targeted interventions in this population (33, 34). For instance, Lorig and colleagues (2001) found that a self-management program targeting self-efficacy in patients with chronic diseases improved health status, self-management behaviors, and healthcare utilization (33).

This study also highlights the effectiveness of a hybrid approach combining telehealth and in-person visits. Telehealth technology improved access to care, reduced barriers, and offered increased flexibility in scheduling provider appointments, but faced challenges such as technological barriers and maintaining rapport between CHAs and participants (15, 35). Adapting to the hybrid model provided personalized care and improved communication while addressing the limitations of CHAP’s initial approach (36). This approach aligns with findings from several studies that have examined the benefits of hybrid interaction models in various populations and contexts. These studies have found that a hybrid telehealth model improved patient satisfaction, access to care, and health outcomes among older adults with multiple chronic health conditions(37–40). Similarly, Hollander & Carr (2020) reported that telehealth and in-person visits enhanced patient engagement and facilitated timely care, particularly during the pandemic when access to healthcare was limited (36). In another study, Greenhalgh and colleagues (2020) emphasized the importance of adaptability and a hybrid approach to ensure personalized, flexible patient care strategies during the pandemic (41). Furthermore, Smith and colleagues (2020) highlighted the benefits of telehealth in expanding access to healthcare and mitigating barriers faced by underserved populations, including older AA adults (15).

Clear communication and robust relationships were vital for the project’s success, as CHAs offered tailored support to older adults (15, 35). The project’s cultural sensitivity enhanced its acceptability among the AA older adult population. A supportive environment for implementation was crucial, with effective communication addressing health disparities and risks associated with the pandemic such as misinformation and vaccine hesitancy (36, 41). Tailoring the intervention delivery ensured compatibility and maintained participant engagement throughout the intervention (29). The current project’s cooperation with local church leaders and FBOs aligns with Smith and colleagues (2014) (2020), who also found that engaging FBOs was crucial in recruiting community health advisors and ensuring the success of their intervention (15). This collaboration reinforces the significance of community engagement and partnership in promoting the reach and impact of health initiatives.

### Limitations

The project took place during the height of the COVID-19 pandemic, which presented unique challenges to managing chronic health conditions for African American older adults, particularly those who were isolated or had limited access to healthcare. Due to the small sample size, statistical comparisons were not feasible.

## Conclusions

Overall, the findings from this study offer significant insights into the implementation process and highlight essential factors to be considered when designing interventions for similar contexts. These insights contribute valuable knowledge about the feasibility, acceptability, and impact of a hybrid (virtual/in-person) lay health advisor model during the pandemic. They can guide the development of future interventions in this domain. Our results are also consistent with existing research on the effectiveness of community-based health interventions for underserved populations. The high completion rate and improvements in chronic health condition management underscore the necessity of incorporating culturally sensitive, contextually relevant, and responsive strategies in healthcare interventions aimed at underserved African American older adults.

## Data Availability

All relevant data are within the manuscript and its Supporting Information files

## Acknowledgements

We would like to extend our sincere appreciation to the following faith-based organizations, all located in South Los Angeles, CA, and their respective leaders for their valuable partnership and support in the completion of this community-centered research project: Beulah Baptist Church (Rev. Dr. Robert L Taylor) Holy Mt. Calvary Missionary Baptist Church (Dr. Leonard White), Mount Salem Missionary Baptist Church (Pastor Patricia Joyce Strong-Fargas), Shiloh Missionary Baptist Church (Dr. Joe Waller), St. Mark Missionary Baptist Church (Dr. Lovely Haynes), and FAME Church (Reverend Judi Wortham). Burning Bush Church (First Lady Lorrie Denson) is in Inland Empire, CA.

## Author Contributions

M.B., S.C. and R.V. were involved in the conception, funding acquisition, and design of the study. M.B., S.B., and E.K.A. performed the data analysis and interpretation. E.K.A., H.S, J.W., and M.B. together drafted the initial manuscript. In addition, E.K.A., H.S., J.W., M.B. and L.W.K. were involved in overseeing the study and data collection. All authors have read and agreed to the published version of the manuscript.

